# Genetic networks suggest Asperger’s Syndrome as a distinct subtype of Autism Spectrum Conditions

**DOI:** 10.1101/2023.09.21.23295892

**Authors:** Adam A. Dmytriw, Sadiq Naveed, Sherief Ghozy, Sara Morsy

**Affiliations:** Neuroendovascular Program, Massachusetts General Hospital & Brigham and Women’s Hospital, Harvard Medical School, Boston MA; Psychiatry Department, University of Connecticut, CT; Frank H. Netter School of Medicine, Quinnipiac University, CT; Infant- Parent Mental Health, University of Massachusetts System, Boston; Department of Neuroradiology, Mayo Clinic, Rochester, MN, USA; Nuffield Department of Primary Care Health Sciences and Department for Continuing Education (EBHC program), University of Oxford, UK; School of Pharmacy and clinical sciences, University of Bradford

**Keywords:** Asperger syndrome, autism spectrum condition, DSM-V, WGCNA, Weighted gene correlation networks

## Abstract

**Background:** The Diagnostic and Statistical Manual of Mental Disorders (DSM-V) issued new diagnostic criteria for autism spectrum conditions (ASC) which excluded cases of Asperger’s syndrome (AS) and pervasive developmental disorder not otherwise specified (PDD-NOS). This negatively affected the support received by those affected. In this study, we explored if AS can be considered a subtype of autism.

**Methods:** We explored if AS is considered a subtype of autism through gene network analysis. We analysed the GEO microarray data of 170 autism patients, then, used a weighted gene co-expression network (WGCNA) pipeline. We explored whether these modules share the same expression patterns across autism subtypes. The study had 170 patients grouped into three groups: AS, autism, and PDD-NOS.

**Results:** Twenty-one genetic modules were constructed for autism spectrum conditions. However, only AS had a significant downregulation in one of the modules. The module genes were significantly involved in the regulation of proteolysis, catabolic process, and NF-kappaB signalling. Pathway analysis showed the association of these genes with RIPK1-mediated necrosis and regulation of necroptotic cell death. Brain enrichment analysis showed a spatiotemporal distribution. There was significant gene enrichment in the prefrontal cortex during infancy and childhood then it changes in adulthood to be more abundant in the cingulum bundle, and corpus callosum.

**Conclusions:** Our results suggest AS can be a distinct subtype of ASD, showing a similar gene expression pattern. Spatiotemporal distribution of the module genes in different brain regions explains the high functionality in individuals with AS.

## Introduction

Autism spectrum conditions are one of the most common neurodevelopmental disorders that is typically diagnosed at a young age. The world health organization (WHO) has estimated that it has a global prevalence of 0.76%-2.67% (Zeidan et al., 2022). Similar to other psychiatric diseases, autism spectrum conditions are diagnosed based on the Diagnostic and Statistical Manual of mental disorders (DSM), a manual produced by the American Psychiatric Association to aid clinicians in diagnosing mental illnesses (Regier et al., 2009). However, changing diagnostic criteria in the DSM-V has resulted in a decreased prevalence of cases due to the exclusion of some cases of pervasive developmental disorder (PDD) and Asperger syndrome based on these criteria (Kulage et al., 2020).

The latest version of the DSM focused on creating dimensions for different mental illnesses rather than categorizing them which reflected the spectrum nature of many psychiatric conditions (Williams & First, 2013). In DSM-V, the four categories of autism: pervasive developmental disorder not otherwise specified (PDD-NOS), Asperger’s disorder, and autistic disorder were combined into one category of Autism. DSM-V excluded Rett’s disorder and childhood disintegrative disorder from the Autism category. Moreover, the DSM-V introduced sensory symptoms as one of the criteria for the diagnosis of repetitive and restricted behaviors (Williams & First, 2013). The DSM-V also added a fifth and new diagnostic category called social communication disorder (SCD), for patients with problems in the social domain only with no restrictive and repetitive behavior (Williams & First, 2013).

DSM-V updates led to the development of a high threshold for diagnosis of ASC, and it resulted in a more accurate estimation of ASC cases (Kulage et al., 2020). However, a study found that the DSM-V had high specificity for autistic spectrum disorders, but a very low sensitivity towards cases of Asperger’s syndrome (McPartland et al., 2012). This means that the new classification will miss cases with Asperger’s syndrome. The study showed that these changes are mainly due to changes in the diagnostic rubric and not, as many anticipated, due to changes in terminologies. The greatest change is arguably the specified list of symptoms for the social communications domain (McPartland et al., 2012).

A recent study assessed the impact of this new classification on persons with Asperger’s syndrome and their families (Smith & Jones, 2020). The study reported that some individuals with Asperger’s syndrome agreed with the new classification as it provided a sense of belonging for all patients on the autism spectrum. The study noticed that this group of patients were comfortable with being identified as autistic. Other persons with AS stated that the new classification affected the identity of persons who were diagnosed with Asperger’s syndrome. These patients were afraid to be stereotyped as disabled persons with, for instance, limited career opportunities (Smith & Jones, 2020).

A study reviewed the current literature and reported that AS is a distinct type of autism as it is significantly associated with co-morbidities (Edelson, 2022). The study found a higher incidence of motor coordination disorders in individuals with Asperger’s syndrome compared to other subtypes. Another study showed AS to be hyperresponsive to sensory stimuli while autism had hypo-responsiveness to auditory and visual stimuli (Edelson, 2022). Another study showed that there is a higher significant risk of schizophrenia, and major depressive disorder in Asperger’s syndrome compared to other subtypes of autism. In addition, there was a high cortical gene correlation between ADHD and AS (González-Peñas et al., 2020).

Despite having high IQs, a study found that children with AS often suffered from social isolation and bullying at an early age due to difficulties in social interactions (Mirkovic & Gérardin, 2019). Many of these individuals still need support and social skills training to improve interpersonal relationships and understand social situations (Mirkovic & Gérardin, 2019). However, the type of training is different than with other ASC individuals specifying the need for the characterization of this subtype in DSM-V (Baghdadli et al., 2013; Mirkovic & Gérardin, 2019).

At the genetic level, individuals with Asperger’s syndrome usually have a family history of autism spectrum conditions compared to other subtypes of autism (Mirkovic & Gérardin, 2019). Specific single nucleotide polymorphisms (SNPs) in the aryl-hydrocarbon receptor nuclear translocator 2 (ARNT2) gene have been identified as a characteristic of individuals with Asperger’s syndrome (Di Napoli et al., 2015). Structural MRI meta-analysis showed distinct grey matter morphometry in AS compared to other types of autism (Kevin et al., 2011).

Despite the currently available body of literature, there has been little investigation into potential neurobiological differences between Asperger’s syndrome and other subtypes of autism. In our study, we investigate the distinct pattern of gene networks in each subtype of autism to explore possible distinctions of a gene signature for each subtype. By expanding our understanding of these distinctions, we can pave the path for more targeted and personalised approaches to diagnosis and therapy in the future.

## Methods

### Participants

The GEOdataset (GSE18123) describes the whole blood gene expression of autistic patients using the Affymetrix array and was therefore used for this study. It consists of two independent datasets (P1 and P2), P1 includes 66 Autism spectrum conditions and 33 control groups (Kong et al., 2012), while the second dataset has 104 autism spectrum patients and 82 control groups. We excluded the control cases as we focused on autism spectrum conditions (ASC). The study used DSM-IV classification for autism, thus, the case groups comprised of autism, Asperger syndrome and Pervasive Developmental Disorder-Not Otherwise Specified (PDD-NOS)

### Statistical analysis

For each dataset, a different platform of Affymetrix microarray profiling was used. Affymetrix array U133p2 was used for the first dataset and the GeneST platform for the second dataset. The data was exported to R statistical package (Team, 2017) using the GEO query package (Davis & Meltzer, 2007).

We processed each dataset individually using the Oligo package (Carvalho & Irizarry, 2010). The steps included background correction, quantile normalization, batch effect removal using the SVA package, filtering, and annotation (Klaus & Reisenauer, 2018). After combining P1 and P2, the expression data was inspected for any duplicates and if there are duplicate genes, the mean of the expression was done. After processing, overlapped genes between two datasets were used to build an expression dataset used for the weighted gene correlation networks (WGCNA) (Langfelder & Horvath, 2008a).

### Construction of a weighted gene correlation network

As previously described, weighted gene co-expression networks were constructed using the WGCNA package (Langfelder & Horvath, 2008b).To identify outlier microarray samples, average linkage agglomerative hierarchical clustering was performed to detect and exclude outlier samples. The steps to construct the gene modules include the construction of an adjacency matrix from Pearson correlation, adjacencies were transformed into topological overlap matrix (TOM), calculation of corresponding dissimilarity (1–TOM), followed by hierarchical clustering using dissimilarity (1–TOM) as the distance measure. Then, a dynamic tree cut algorithm with a module size of 20 and a minimum cut height of 0.95. The gene module is a collection of genes with high topological overlap similarity. Module eigengene is the first principal component of gene expression. To identify genes that construct a module, module membership is calculated as the association between a gene and module eigengene of a given module (MM). For each gene in the module, we calculate the gene significance (GS) which is the absolute value of correlation between gene expression values and each trait. The hub gene is defined as the gene with high GS, MM and high intramodular connectivity (Langfelder & Horvath, 2008a). After the construction of the gene modules, correlated the occurrence of each subtype of ASC to the module eigengene.

### Enrichment analysis

We performed gene enrichment analysis for genes of significant modules. A gene ontology analysis (GO) was used to identify characteristic cellular components, biological processes and molecular functions. Brain enrichment analysis was performed using the ABA enrichment package that uses Allen Brain Atlas data (Grote et al., 2016). The package does gene set expression enrichment in the adult and developing human brain. After getting the enrichment data, we used the ggseg package for brain enrichment visualization (Mowinckel & Vidal-Piñeiro, 2020).

## Results

### Patient characteristics

170 patients were split into three categories: those with Asperger’s syndrome (n = 24), those with autism (n = 72), and those with Pervasive Developmental Disorder-Not Otherwise Specified (PDD-NOS) (n = 74). Six patients were omitted from the analysis: one patient with autism had Landau Kleffner Syndrome Table 1. Two autistic patients have 16p13.1 amplification and deletion 11p11.12; one PDD-NOS patient has 16p13.1 amplification; and one case of Asperger’s syndrome has 3p duplication, which is maternally inherited. In addition, 7q21 amplification, 3q26.327 amplification, and 16p13.1 amplification were found in three PDD-NOS cases.

**Table 1.**
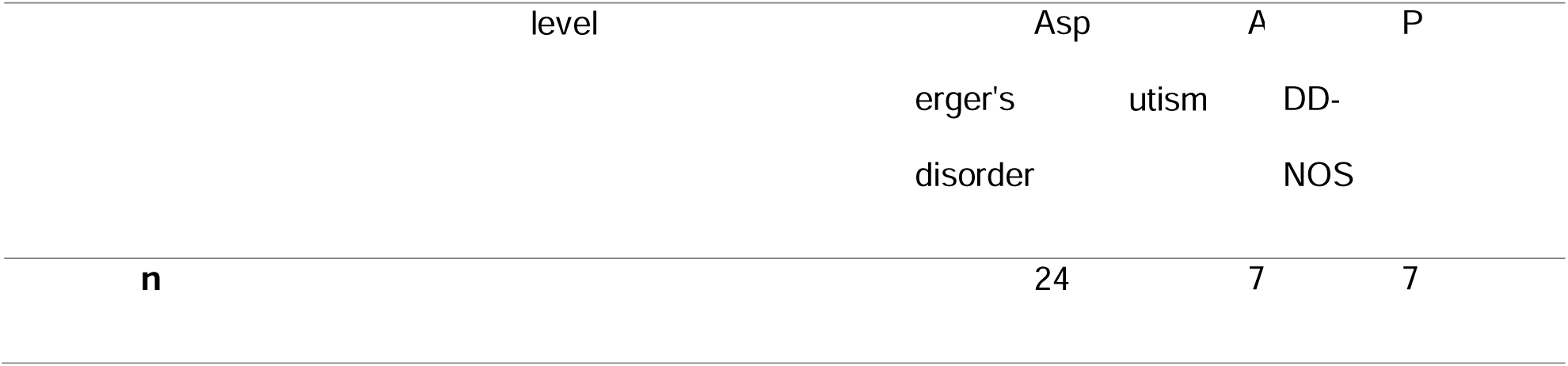
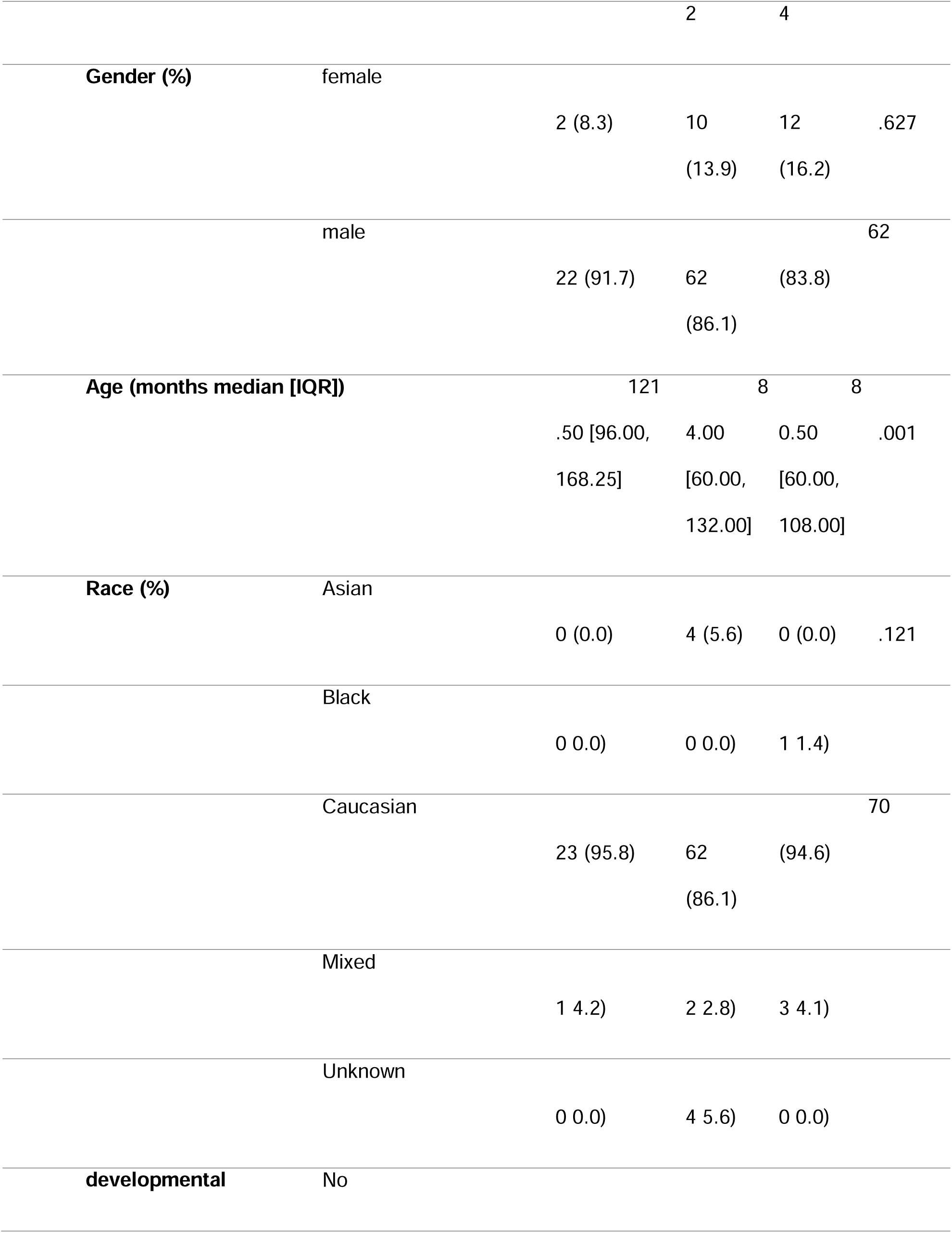
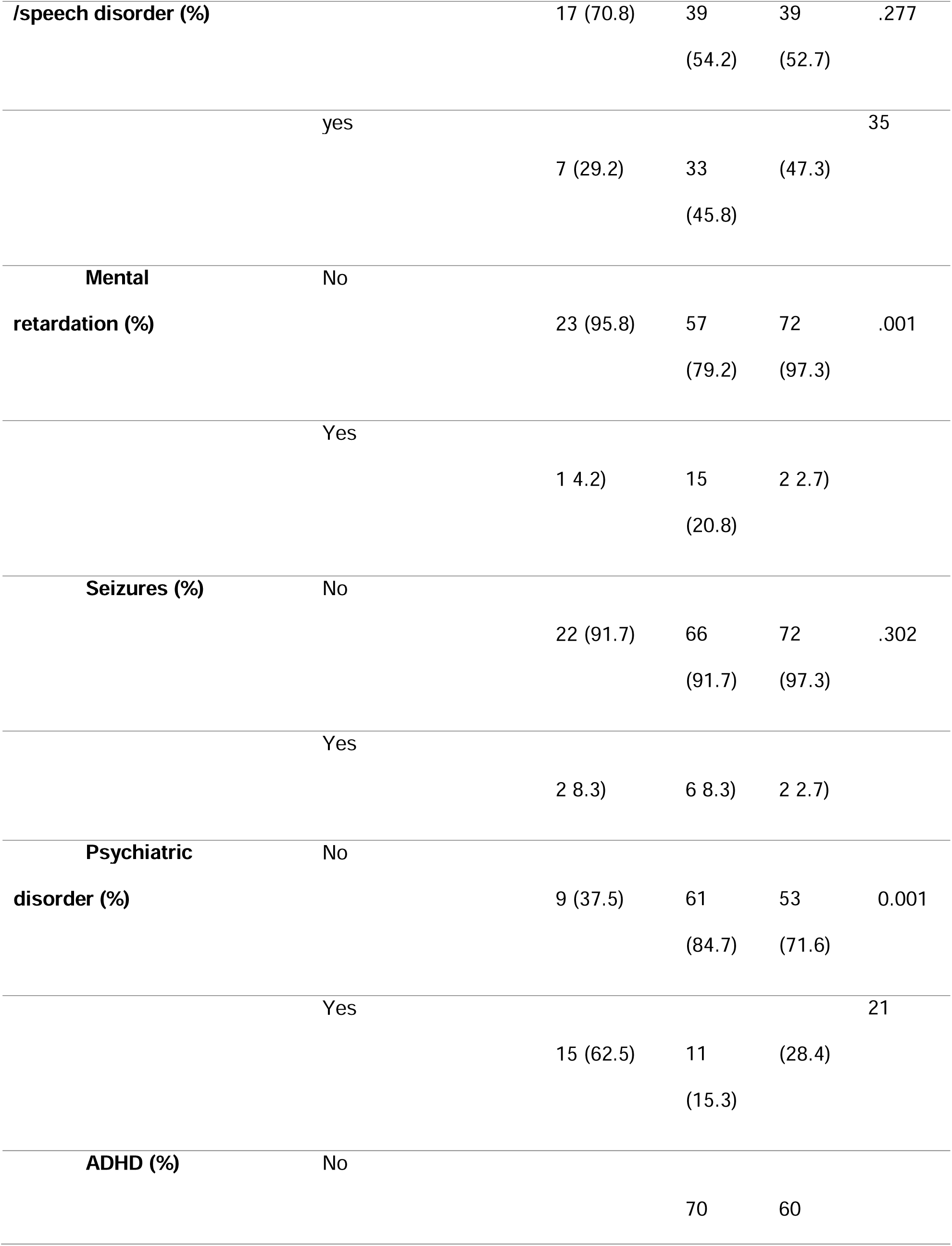
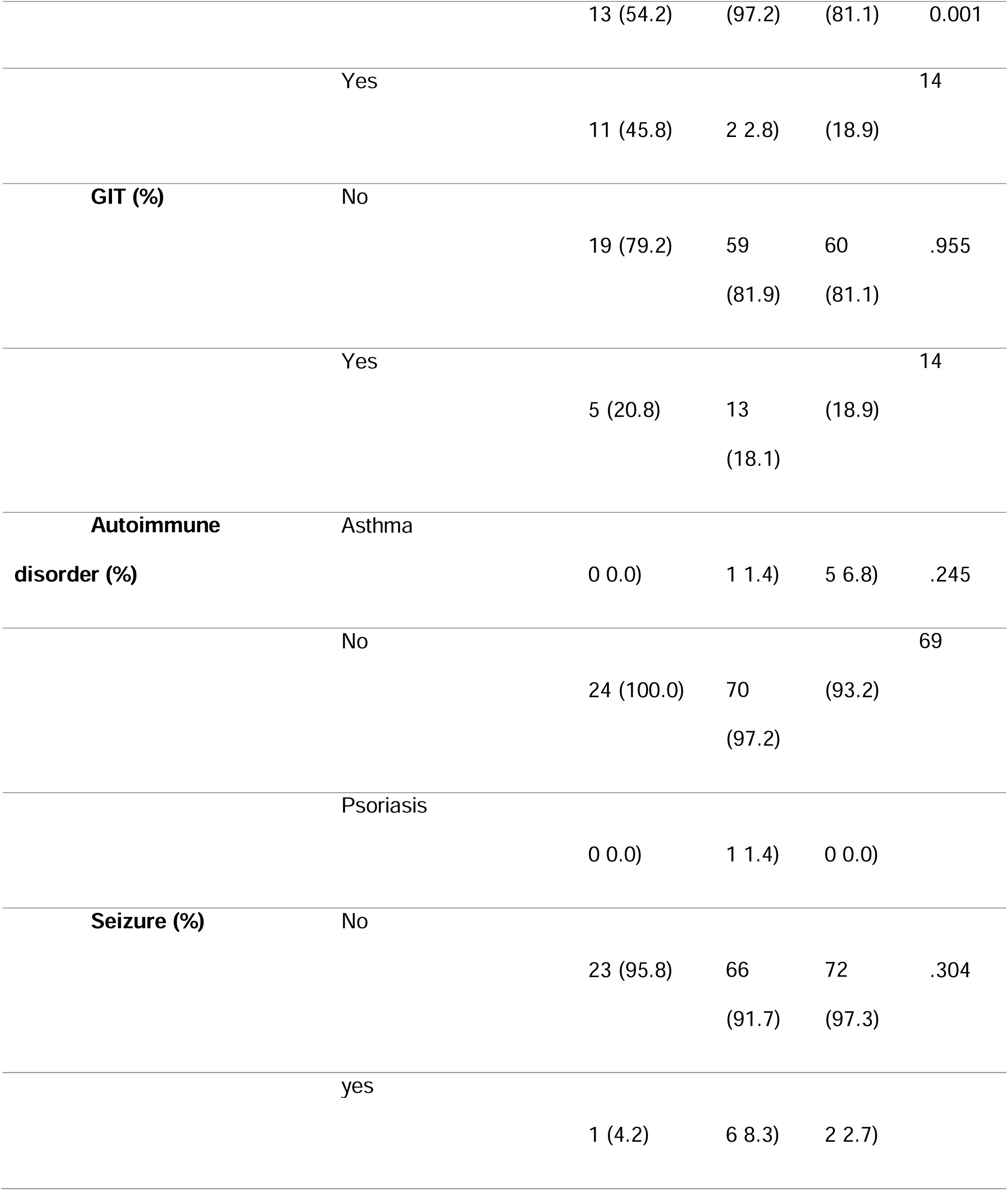
summarizes the characteristics of individuals diagnosed with three subtypes of autism spectrum condition, as per the Diagnostic and Statistical Manual of Mental Disorders, Fourth Edition (DSM-IV)

Co-morbidities were most predominant in patients with autism Table 1. Post-hoc pairwise comparisons showed that the number of patients with psychiatric co-morbidities is significantly higher in Asperger’s syndrome compared to autism but significantly lower than PDD-NOS. However, there was no significant difference between autism and PDD-NOS. For intellectual disability, there was a significant difference between the three groups, however, pairwise post hoc analysis showed only a significantly higher number of patients with intellectual disability in autism compared to PDD-NOS and Asperger’s syndrome. For ADHD, there was a significantly higher number of ADHD in Asperger’s syndrome and PDD-NOS than in autism.

### Comparing Genetic modules for three types of autism spectrum conditions

The WGCNA yielded 21 modules of co-expressed genes in autism. We found that there is only significant decrease of gene expression in one module in patients with Asperger’s syndrome compared to PDD-NOS and Autism. However, no significant module enrichment in any of the other categories against each other and autism. The green module was negatively enriched in Asperger’s disorder compared to autism and PDD-NOS indicating distinct genetic signature Figure 1.

**Figure 1.**
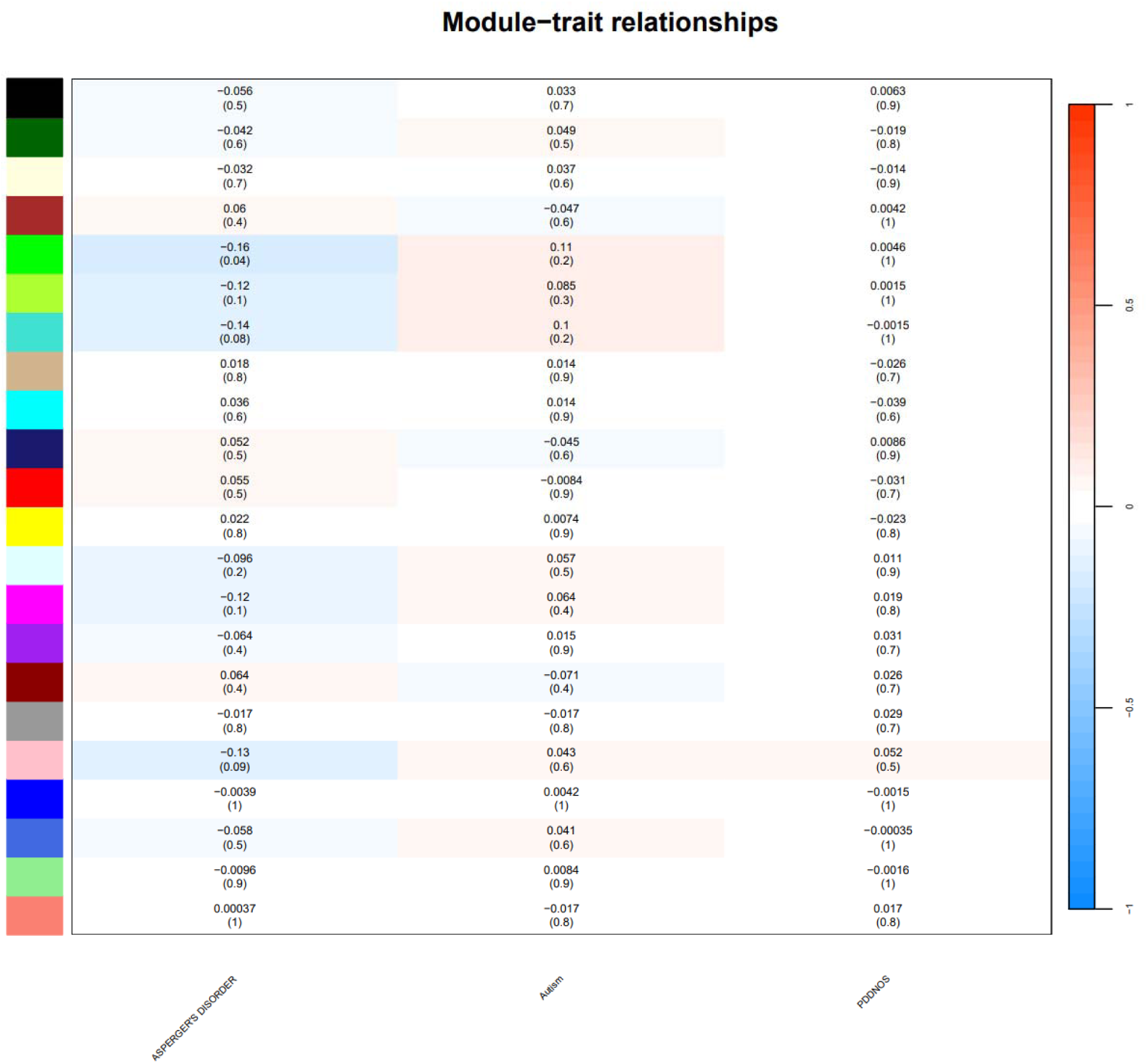
showing the gene module and the corresponding correlation between the occurrence of each subtype of ASC versus other types. The values represent correlation value and *p-value. Each network of genes is identified by colour. The green module was the only module that was downregulated in Asperger’s syndrome*.

### Transcriptomic profile of the green module

The green module had 280 genes that were negatively regulated in Asperger’s syndrome compared to other autism subtypes. The Hub gene was CYB5R4 Figure 2.

**Figure 2.**
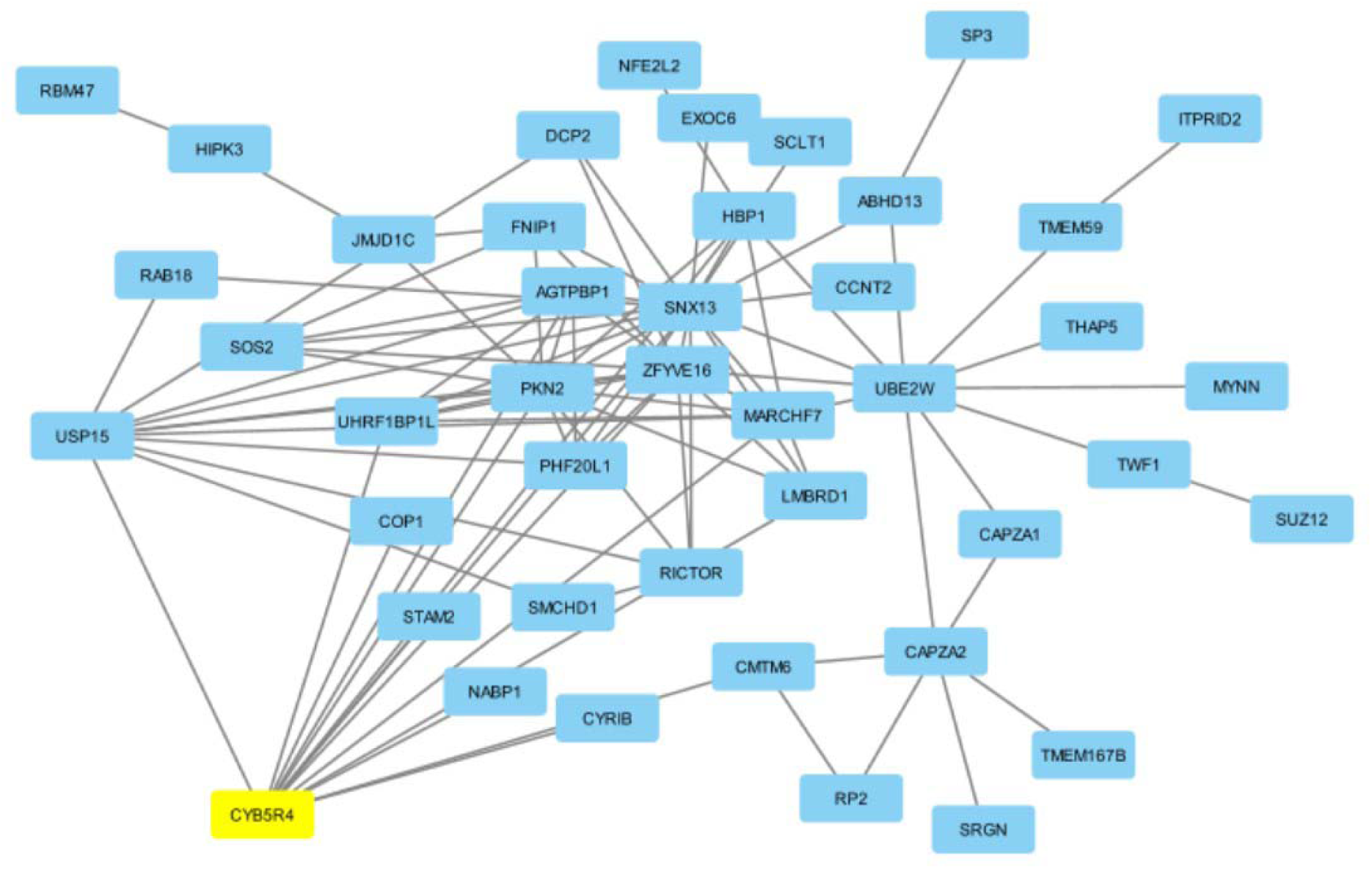
Gene network of the top hub genes in the green module and shows that CYB5R4 as the main hub gene.

Gene ontology enrichment analysis showed that the green module gene is enriched in genes associated with proteasome-mediated ubiquitin-dependent protein catabolic process, positive regulation of proteolysis and NF-kappaB signalling. The genes coded for proteins that are part of secretory granule membrane, lysosomal membrane, early endosome and nuclear envelope. The gene network was involved in metabolic processes associated with the binding with GDP and 1-Phosphatidylinositol binding Figure 3.

**Figure 3.**
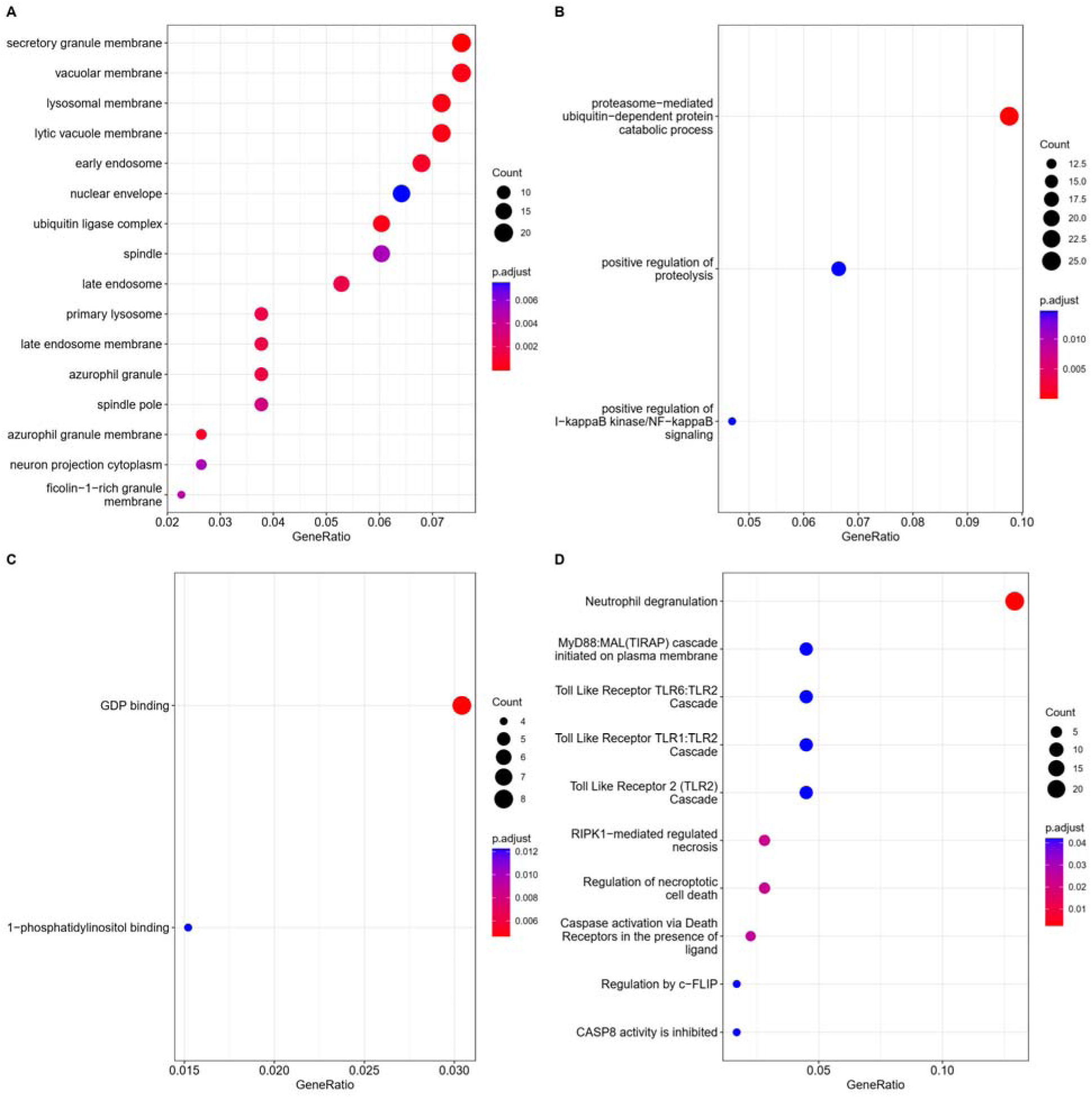
Gene ontology annotation for the green module genes representing enrichment analysis for A) Cellular components B) Biologic processes C) Molecular functions D) Reactome pathway enrichment analysis.

Reactome pathway analysis showed that 20 of these genes were involved in neutrophil degranulation; 5-10 genes were involved with MyD88 adapter-like (Mal) (TIRAP) cascade initiated on the plasma membrane and different Toll-Like receptor cascade, RIPK1-mediated regulated necrosis and regulation of necroptotic cell death Figure 3. Through the enrichr package, we identified key transcription factors regulating the module gene expression which were SOX4, JARID2 and TP53 Figure 4.

**Figure 4.**
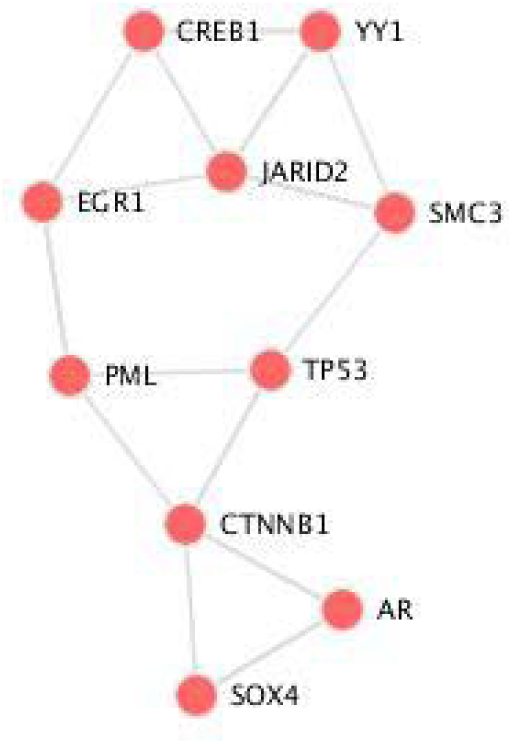
Network of transcription factors that control gene expression in the green module

Brain enrichment analysis across different age groups showed a temporal pattern of enrichment for the green module genes. During infancy, the genes were enriched in the prefrontal cortex, mostly in the ventrolateral prefrontal cortex, and posteroventral (inferior) parietal cortex. Then in childhood, these genes become more abundant in the posteroventral (inferior) parietal cortex, primary auditory cortex (core), and ventrolateral prefrontal cortex Figure 5.

**Figure 5.**
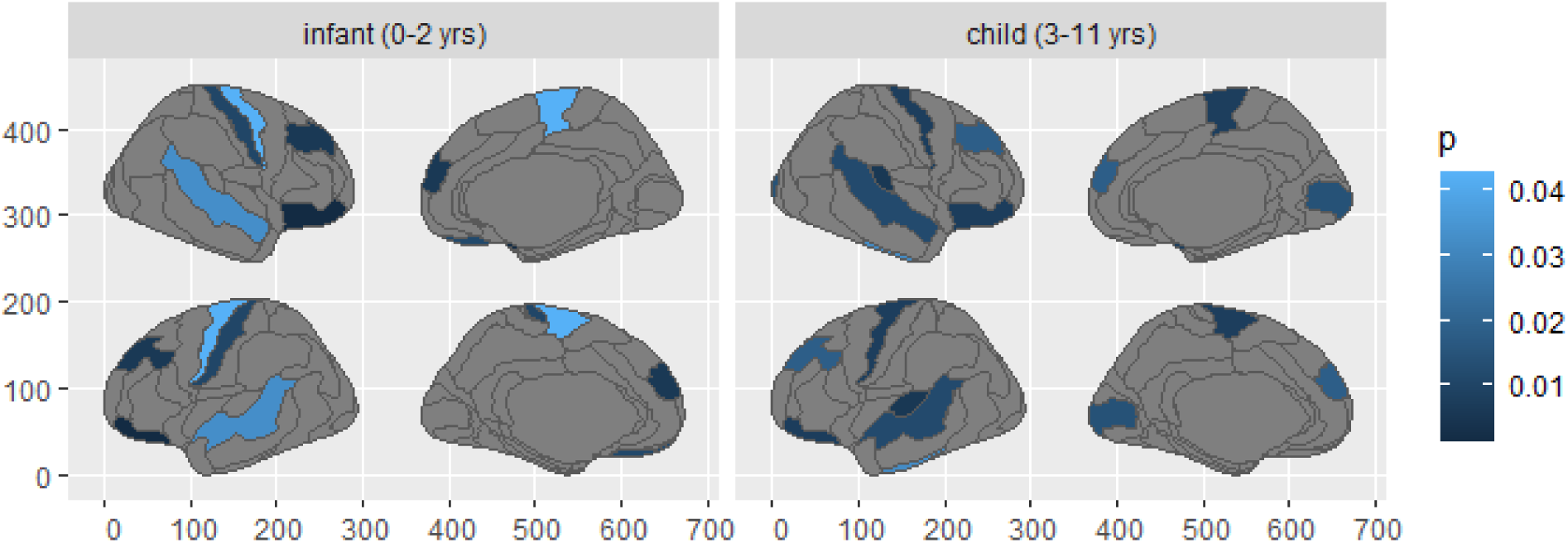
Gene enrichment analysis in right and left hemisphere in infants and children, the analysis shows the genes were enriched in the prefrontal cortex, mostly in the ventrolateral prefrontal cortex, and posteroventral (inferior) parietal cortex in infants then it changes to be enriched in the posteroventral (inferior) parietal cortex, primary auditory cortex (core), and ventrolateral prefrontal cortex.

In adults, there is a complete change in the gene enrichment pattern to enriched in the corpus callosum and right cingulum bundle in the adult brain Figure 6.

**Figure 6.**
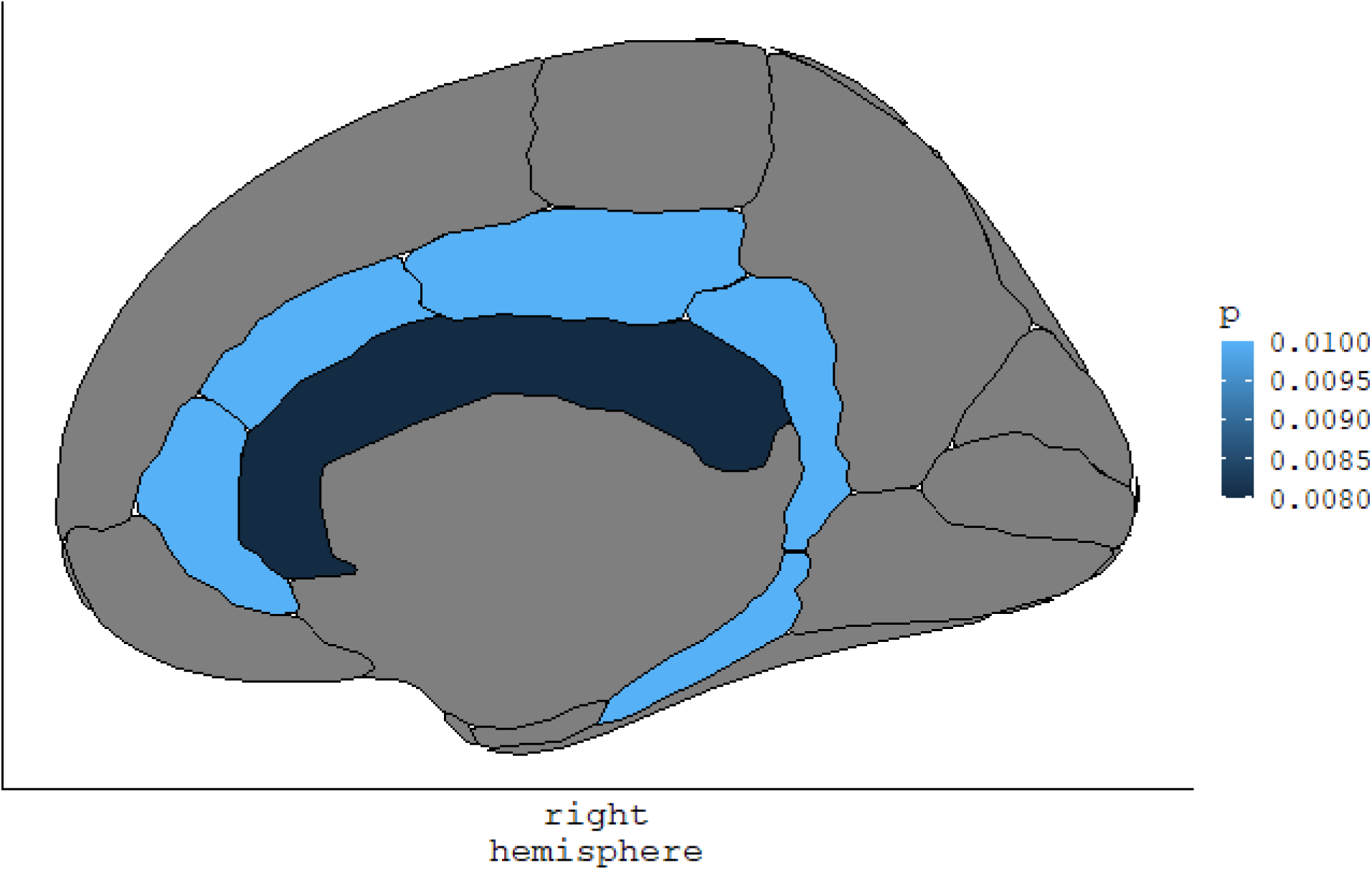
right brain hemisphere showing gene enrichment in adults; as in the figure, the gene are mainly enriched in corpus callosum and cingulum bundle.

## Discussion

Our results show a genetic distinction in Asperger’s syndrome compared to other subtypes of autism spectrum conditions. In this cohort of patients, we found that there is a specific gene network that was negatively expressed compared to other subtypes of autism spectrum conditions. The gene network is enriched in genes responsible for different catabolic processes including proteolysis and different metabolic molecules. Interestingly, brain enrichment analysis showed that these genes had spatiotemporal enrichment. The gene expression levels change from the prefrontal cortex, mostly in the ventrolateral prefrontal cortex, and posteroventral (inferior) parietal cortex in infancy to become more abundant in posteroventral (inferior) parietal cortex, primary auditory cortex (core), and ventrolateral prefrontal cortex during childhood. In adults, there is a complete change of the gene enrichment pattern to be more abundant in the corpus callosum and right cingulum bundle in the adult brain.

The hub genes of the green module were cytosolic NADH cytochrome b5 oxidoreductase (NCB5OR) or CYB5R4 gene which codes for functional domains of cytochrome b5 and cytochrome b5 reductase. The loss of gene expression of CYB5R4 in the cerebellum and midbrain was associated with altered proprioception, and locomotor abnormalities especially in iron-deficient mice (Stroh et al., 2016). In addition, another study found that mutation in CYB5R4 led to an abnormal version of NCB5OR that was found in Japanese patients diagnosed with epilepsy, trigonocephaly and pervasive developmental disorders of autism. Further investigations showed that these abnormalities in gene expression were associated with epilepsy and/or craniofacial abnormalities (Suzuki et al., 2020). Worth mentioning that a western Australian population-based study showed that craniofacial abnormalities are associated with high incidence of Autism spectrum conditions(Junaid et al., 2022). The module is associated with decreased catabolic process for protein. Interestingly, Stroh et al. found that loss of NCB5OR in the cerebellum disturbs iron dyshomeostasis as evident by decreased catabolism of iron regulatory protein 2 and metallothionein2 (Stroh et al., 2016). However, more studies are needed to explore how it is more linked to Asperger’s syndrome compared to other subtypes.

Gene enrichment analysis showed that these genes are associated with proteolysis, protein catabolic process and Nuclear Factor Kappa B (NF-kB) signalling. The genes were specifically abundant in the lysosomal membrane and secretory granule membrane.

Research showed that autistic patients share defects in synaptic development mainly due to increased brain-derived neurotrophic factor (BDNF) which is mainly due to a defect in the proteolytic process of pro-BDNF which leads to abnormal connectivity and synaptic plasticity causing changes in behaviour (Han et al., 2022; Koh et al., 2014). Moreover, Ormstad et al. showed that increased tryptophan and decreased synthesis of 5 hydroxy tryptophan is the hall mark for Asperger’s syndrome compared to other subtypes of autism (Ormstad et al., 2018). Another study that was performed on Asperger’s syndrome and autistic children revealed there is a defect in the proteolysis of casomorphins (CM) that led to higher levels of urine CM-7 (Sokolov et al., 2014). Interestingly, the severity of clinical symptoms was positively correlated with the level of urinary CM-7 (Sokolov et al., 2014). This pattern was not reported in other subtypes of autism spectrum conditions.

The transcriptomic profile of the gene network that is distinctively downregulated in Asperger’s syndrome showed enrichment for proteasome-mediated ubiquitin-dependent protein catabolic process, positive regulation of proteolysis and NF-kappaB signalling. The gene network was involved in metabolic processes associated with the binding with GDP and 1-phosphatidylinositol binding.

Studies have shown Asperger’s syndrome and autism patients had altered proteolysis of mitochondrial IMMP2L that was linked to social behavioural changes (Bertelsen et al., 2014; Petek et al., 2007). Another interesting biologic process ubiquitin-dependent protein catabolic process. A study conducted on Chinese patients showed that aberrantly expressed miR-103a-3p explained the abnormal ubiquitin-mediated proteolysis in children diagnosed with Asperger’s syndrome (Huang et al., 2015). Similar protein dysregulations patterns were also evident in individuals with Asperger’s syndrome compared to other subtypes of autism spectrum conditions (Steeb et al., 2014). Our results also showed that module genes were also involved in NF-kB signalling. A systematic review showed that NF-kB was dysregulated in all individuals with autism explaining the mitochondrial dysfunction and neuroinflammatory response (Liao & Li, 2020). These genes were involved in GDP and 1-phosphatidyl inositol binding. Both molecules are essential for different metabolic processes including the synthesis of 3-phosphorylated inositol lipids. A study showed that autism was associated with abnormalities in polyunsaturated fatty acid metabolism (Das, 2013), however, no specific study investigated whether there is characteristic metabolic signature for autism.

Our results also showed that the green module was mainly regulated by three transcription factors SOX4, JARID2 and TP53. SOX4 is a member of the SRY-related High Mobility Group (HMG) box or SOX family of transcription factors. SOX4 is a transcription factor that is essential for the regulation of stem cell differentiation and progenitor development. It is involved in central and peripheral nervous system development (Angelozzi & Lefebvre, 2019). SOX 4 has been involved in the development of intellectual disability syndrome with mild facial dysmorphism (Zawerton et al., 2019). Another study showed that SOX4 haploinsufficiency was associated with non-specific neurodevelopmental disorders including the abnormal development of cognitive function (Marco et al., 2022) which can be responsible for specific cognitive characteristics in Asperger’s syndrome.

One of the theories for the development of autism spectrum conditions is immune imbalance. A study found that immune function genes like CD99L2, JARID2 and TPO were associated with the development of autism spectrum conditions (Ramos et al., 2012). Jumonji and AT-rich interaction do-main 2 (JARID2) gene encode ARID transcription factor which is included in embryonic development and maintenance of cell type identity (Ramos et al., 2012; Verberne et al., 2021; Viitasalo et al., 2022). JARID2 haploinsufficiency was associated with distinct neurodevelopmental disability and autistic features (Verberne et al., 2021; Viitasalo et al., 2022). Our results showed that JARID2 is one of the main transcription factors that are connected to the green module. A study showed that Asperger’s syndrome is associated with immune dysfunction as evidenced by high serum IGE and atopy (Magalhães et al., 2009).

Our results showed spatial and temporal brain enrichment of the genes in Asperger’s syndrome. During infancy and childhood, the genes were significantly expressed in the prefrontal cortex and posteroventral parietal cortex. In childhood, there was also significant expression in the primary auditory cortex. Literature showed that there is an increased number of neurons in the prefrontal cortex in children. White matter abnormality has been associated with the development of autism. Ikuta et al. showed that the development pattern of cingulate matter that occurs in late adolescence and early adulthood was associated with executive dysfunction in autism spectrum conditions (Ikuta et al., 2014). Another longitudinal study showed that there was slower development of fractional anisotropy in the cingulum bundle, superior longitudinal fasciculus, internal capsule, and splenium of the corpus callosum (Andrews et al., 2021). Moreover, the severity of autism during child development was dependent on the rate of development of these brain regions. These results coincide with the pattern of gene enrichment for Asperger’s syndrome indicating that AS needs to be reexamined as one of the subtypes of autism (Andrews et al., 2021).

The green module genes were enriched during adulthood in the corpus callosum, Para hippocampus and right cingulate gyrus. A study showed the same in autism spectrum condition, as well as decreased metabolism in these areas (Chien et al., 2021). Another study found that autistic patients share the same behavioural characteristic as adults with callosal agenesis (Paul et al., 2014).

In terms of comorbidities, our study found that compared to other subtypes of autism spectrum conditions, children with Asperger’s syndrome have higher incidence of ADHD and other psychiatric diseases. This was in line with other research studies that showed not only that it had higher incidence of these diseases in patients with Asperger’s syndrome but also similar gene expression pattern (González-Peñas et al., 2020).

## Conclusion

Our analysis shows that Asperger’s syndrome is biologically distinct from other types of autism spectrum conditions. The study elaborates that transcriptomics profiles of children with Asperger’s syndrome showed spatiotemporal enrichment in the brain that might be related to the development of their cognitive function as they grow older. More studies are needed to see how the biological difference between Asperger’s syndrome and other types of autism will inform the care these children receive.

## Data Availability

All data produced are available online at GEO database (ID: GSE18123).

## Data statement

We used the microarray data that is publicly available through GEO database (ID: GSE18123)

